# Genetic variation in *RCOR1* is associated with tinnitus in UK Biobank

**DOI:** 10.1101/2020.09.11.20192583

**Authors:** Helena R.R. Wells, Fatin N. Zainul Abidin, Maxim B. Freidin, Frances M.K. Williams, Sally J. Dawson

## Abstract

Tinnitus is a prevalent condition in which perception of sound occurs without an external stimulus. It is often associated with pre-existing hearing loss or noise-induced damage to the auditory system. In some individuals it occurs frequently or even continuously and leads to considerable distress and difficulty sleeping. There is little knowledge of the molecular mechanisms involved in tinnitus which has hindered the development of treatments. Evidence suggests that tinnitus has a heritable component although previous genetic studies have not established specific risk factors. We performed a case-control genome-wide association study for self-reported tinnitus in 172,608 UK Biobank volunteers. Three variants in close proximity to the *RCOR1* gene reached genome wide significance: rs4906228 (p=1.7E-08), rs4900545 (p=1.8E-08) and 14:103042287_CT_C (p=3.50E-08). *RCOR1* encodes REST Corepressor 1, a component of a co-repressor complex involved in repressing neuronal gene expression in non-neuronal cells. Eleven other independent genetic loci reached a suggestive significance threshold of p<1E-06.

## Introduction

Tinnitus, often referred to as “ringing in the ears” has a reported prevalence of 10-15% in the adult population^1-5^ although the diagnosis and definition of tinnitus remains inconsistent. For most individuals, tinnitus is short lived and not unduly problematic but in some individuals it can be long-lasting and frequent leading to considerable distress and anxiety with 0.5% of individuals reporting it severely affects their ability to live a normal life^3^. Individual differences in the presentation, duration and frequency of these phantom sounds suggests that tinnitus is likely to be a heterogeneous condition representing a range of pathologies. It often manifests as secondary to hearing loss in both permanent hearing loss or temporary loss secondary to recreational or occupational noise exposure^1-7^. Although most individuals with tinnitus also have a hearing loss, tinnitus can occur in isolation^2; 3^ The relative contributions of the central and peripheral auditory system to tinnitus remain uncertain but evidence suggests that acute temporary tinnitus after noise exposure is likely to reflect cochlear damage while long term chronic tinnitus has been shown to involve re-organisation or disturbance of neurons in the primary auditory cortex owing to a lack of signal from the cochlea^1; 8^. Lack of understanding of the mechanisms involved have meant that current therapies to address tinnitus symptoms are limited to masking by external sound sources or strategies designed to help with the anxiety and stress caused by tinnitus.

Genetic analysis using linkage or genome wide association studies (GWAS) can be a powerful tool to reveal underlying causes in heritable conditions. Evidence that tinnitus has a sizable genetic component has been mixed^9; 10^. There is little evidence to support familial segregation of tinnitus except in rare cases but a recent heritability estimate from more than 70,000 twins in the Swedish Twin Registry suggested that tinnitus heritability at 0.43^11^. This rises to a relatively high rate of 0.68 when the trait is limited to bilateral tinnitus in men only^11^, suggesting a higher genetic component in some. Another Swedish study using a large cohort of adoptees also estimated tinnitus to have a similar value of 0.38 for the heritability for tinnitus^12^. Whether any genetic component to a tinnitus phenotype is specific or may instead represent a secondary phenotype due to increased susceptibility to noise induced damage to the auditory system, remains to be clarified. Identification of the genetic variants involved in tinnitus would help reveal the nature of the mechanisms involved in generating tinnitus after hearing loss, a pre-requisite for development of treatments. Previous pilot genome-wide association studies and candidate gene studies for tinnitus^10; 13-18^ have lacked sufficient power to establish specific genetic risk factors but the relatively high heritability demonstrates there is potential to use such approaches to reveal the underlying mechanisms^9; 10^. On this basis we performed a genome-wide association study using self-reported tinnitus available from 172,608 UK Biobank (UKBB)^19^ volunteers.

## Results

### Phenotype Definition

In the UKBB 172,608 participants responded to the question regarding whether, and how often, they had experienced tinnitus symptoms (Table 1). The 48,728 who had responded yes to experiencing tinnitus answered an additional question regarding how much they were affected by their tinnitus (Table 2). Almost a third of the sample reported experiencing symptoms of tinnitus either at the time of data collection or in the past (Table 1). Of the subset that had experienced or currently experience symptoms, 3.55% reported that symptoms “severely worry, annoy or upset” them when they are at their worst, while a third of these participants’ reported that they are “not at all bothered” by the symptoms. For the genome-wide association study cases were assigned as participants who answered ‘yes’ tinnitus was present a lot, most, or all of the time (N= 14,829); just under 9% of respondents. We chose only these respondents as cases in order to limit our analysis to those individuals who reported frequent tinnitus to improve genetic power since there is evidence of higher heritability in more severe tinnitus subtypes^11^. Controls were assigned to those who have never experienced tinnitus (N=119,600), around 71% of the sample. Replies to the question of how much participants were affected by their tinnitus were not used for phenotype definition as there is a greater degree of subjectivity to this experience. For the GWAS, samples were then further selected based on ethnicity and additional quality control measures (see Methods for details) which resulted in a final sample size of N=91,424 for association analysis.

**Table 1.** Prevalence of tinnitus in UK Biobank participants. Percentage values correspond to the proportion of participants that selected one of these five responses (N=168,328 participants). An additional 4,280 participants selected either “Do not know” or “Prefer not to answer”. %M and %F is the percentage answers for males and females respectively. Case Control Definition designates if the participants who gave these responses were included in the GWAS as Cases or Controls.

| Response | N | All % | % M | % F | Case Control Definition |
| --- | --- | --- | --- | --- | --- |
| **Yes, now most or all of the time** | **10,719** | **6.37** | **8.29** | **4.76** | **Case** |
| **Yes, now a lot of the time** | **4,110** | **2.44** | **2.88** | **2.08** | **Case** |
| Yes, now some of the time | 15,031 | 8.92 | 9.41 | 8.53 | - |
| Yes, not now, but have in the past | 18,868 | 11.21 | 11.13 | 11.28 | - |
| Sub-total who answered Yes | 48,728 | 28.95 | 31.71 | 26.65 | - |
| **No, never** | **119,600** | **71.05** | **68.29** | **73.36** | **Control** |

**Table 2.** Self-reported effect of tinnitus on UKBB participants who reported tinnitus (n=48,728). Percentage values correspond to the proportion of participants that selected one of these four responses. An additional 424 participants selected either “Do not know” or “Prefer not to answer”.

| Response | N | % |
| --- | --- | --- |
| Severely | 1,714 | 3.55 |
| Moderately | 8,037 | 16.64 |
| Slightly | 23,098 | 47.82 |
| Not at all | 15,455 | 31.99 |

### Genome-wide Association Analysis

A linear mixed-effects model was used to test for association between 9,740,198 SNPs and tinnitus, using BOLT-LMM v.2.2^20^. Three SNPs exceeded genome-wide significance (P<5E-08) at the same locus on chromosome 14 (Figures 1 and 2; Supp Table 1). An additional 88 SNPs were associated at a suggestive level of P<1E-06 (Supp Table 1). Conditional and joint analysis using GCTA-COJO^21^ suggested they represent a single genome-wide significant locus and eleven independent suggestive loci (Table 3).

**Figure 1.**
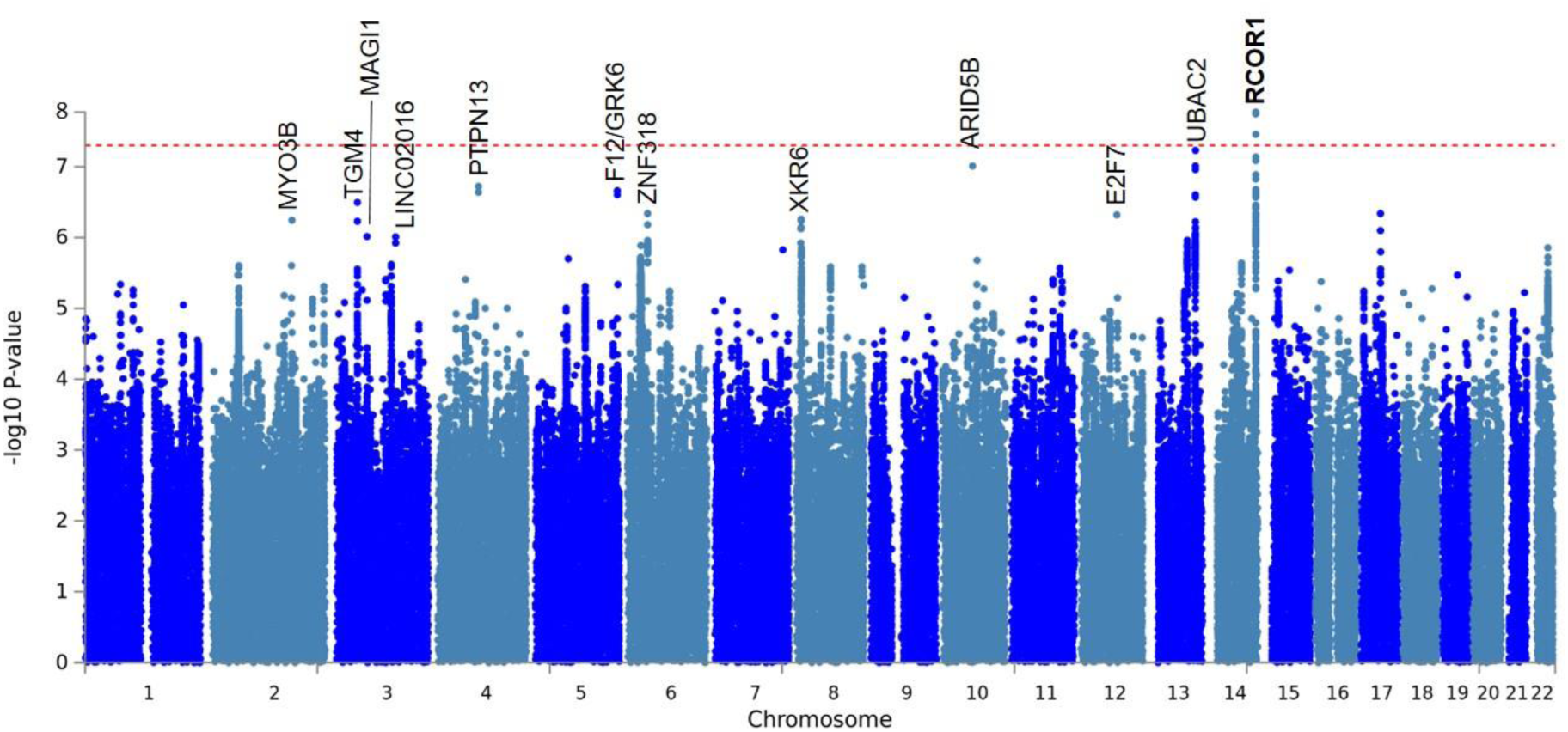
Manhattan plot for genetic association analysis with tinnitus symptoms. The red dotted line marks the genome-wide significance threshold of P<5E-08. Gene loci with p values less than p<1 ×10^-6^ are labelled. The genome wide significant locus is shown in bold.

**Figure 2.**
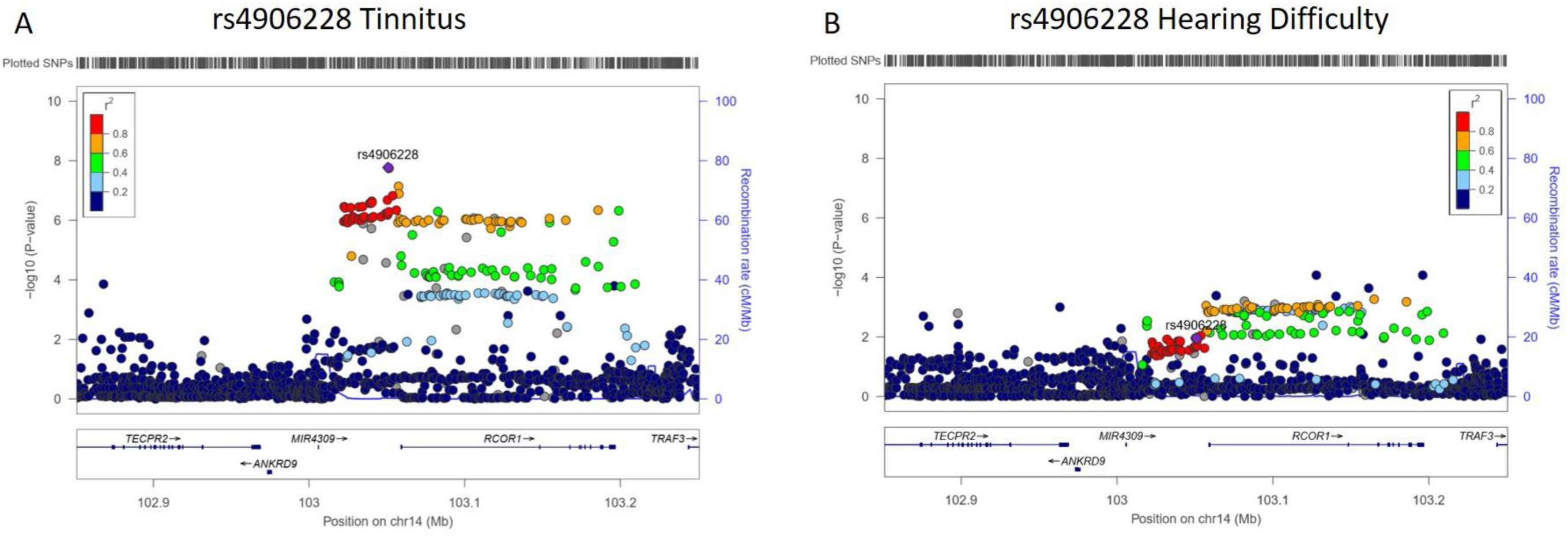
Locus Plots for rs4906228 association with tinnitus and hearing difficulty in UKBB. (A) Locus plot for lead SNP at genome-wide significance on Chr14, rs4906228 in genetic association analysis with frequency of tinnitus symptoms. Purple indicates lead independent SNP generated from GCTA-COJO conditional analysis. The colouring of remaining SNPs is based on the linkage disequilibrium (r^2^) with the leads. Where LD information is not available, SNPs are coloured grey. The genes within the region are annotated and the direction of transcription is indicated by arrows. (B) Same plot is shown for association of rs4906228 with self-reported hearing difficulty as described previously^22^.

SNP heritability estimates for tinnitus calculated using BOLT-LMM gave h2g=0.105, SE=0.003. The estimate was then recalculated to the liability scale, using a tinnitus prevalence derived from the cases and controls above of 0.125 for tinnitus to give a heritability, h2g=0.30. The LD score regression intercept for the tinnitus association analysis was 1.0003, suggestive of a very minor type 1 error rate inflation. The ratio (intercept-1)/(mean(χ2)-1), was 0.0019, which represents the proportion of inflation in the χ2 statistic that the intercept attributes to alternative explanations than polygenicity.

**Table 3.** Summary statistics for lead SNPs that reached a significance threshold of p<1E-06, ordered by p value. Bold denotes genome-wide significance at p<5E-08. Chr, chromosome; SNP, lead SNP; bp, base position; refA, reference allele; beta, effect size from BOLT-LMM approximation to infinitesimal mixed model; se, standard error of the beta; p-value, non-infinitesimal mixed model association test p-value, pJ, joint p-value for variation at loci calculated with GCTA-COJO; Gene, denotes the closest protein coding gene to the variant; Distance, is the distance from the gene (b.p.); Context denotes the consequence of the variant identified by VEP. *denotes genes also identified in previous UKBB GWAS of self-reported hearing^22^. ^ rs72960531 is in an intergenic region without protein coding genes but is within an intron of the lincRNA *LINC02016*.

| Chr | SNP | refA | beta | se | p | pJ | Gene | Distance | Context |
| --- | --- | --- | --- | --- | --- | --- | --- | --- | --- |
| **14** | **rs4906228** | **A** | **0.080** | **0.014** | **1.70E-08** | **1.74E-08** | ***RCOR1*** | **8234** | **Upstream** |
| 13 | rs7336872 | T | 0.072 | 0.013 | 5.90E-08 | 5.94E-08 | *UBAC2* | 0 | Intron |
| 10 | rs118053011 | C | -0.127 | 0.024 | 9.80E-08 | 9.89E-08 | *ARID5B\** | 6901 | Downstream |
| 4 | rs113655471 | T | -0.188 | 0.036 | 1.90E-07 | 1.87E-07 | *PTPN13* | 0 | Intron |
| 5 | rs17876046 | G | -0.288 | 0.056 | 2.20E-07 | 2.18E-07 | *F12/GRK6* | 0 | Intron |
| 3 | rs1532898 | A | -0.069 | 0.013 | 3.20E-07 | 3.24E-07 | *TGM4* | 0 | Intron |
| 6 | rs553448379 | T | -0.063 | 0.013 | 4.60E-07 | 4.58E-07 | *ZNF318\** | 10571 | Downstream |
| 12 | rs7314493 | A | -0.067 | 0.013 | 4.80E-07 | 4.84E-07 | *E2F7* | 0 | Intron |
| 8 | rs4370496 | G | -0.070 | 0.014 | 5.50E-07 | 5.48E-07 | *XKR6* | 0 | Intron |
| 2 | 2:171146084_CTT_C | CTT | -0.283 | 0.057 | 5.70E-07 | 5.69E-07 | *MYO3B* | 0 | Intron |
| 3 | rs557511691 | C | 0.065 | 0.013 | 9.70E-07 | 9.77E-07 | *MAGI1* | 0 | Intron |
| 3 | rs72960531 | C | 0.125 | 0.026 | 9.80E-07 | 9.80E-07 | *LINC02016*<sup>^</sup> | - | Intergenic |

The Variant Effect Predictor (VEP)^23^ was used to map independent lead SNPs to the nearest protein coding genes, using the GRCh37 genomic reference. Summary statistics and gene annotation for the lead SNP at each of these regions are presented in Table 3. The genome-wide significant lead SNP rs4906228 lies upstream of the *RCOR1* gene which encodes REST Corepressor 1, a component of a transcriptional repressor complex which represses neuronal gene expression in non-neuronal cells^24^. Of the 12 independent loci having p<1E-06, eight variants lie within gene introns and three more are positioned upstream or downstream of protein coding genes. Only variant rs72960531 does not lie within or proximal to a protein coding gene, this variant is within intron 3 of a long non-coding RNA (lncRNA), *LINC02016*.

Two of the 12 gene loci were also associated with self-reported hearing difficulty in our previous GWAS in UKBB; *ZNF318* and *ARID5B*^22^. In order to investigate whether tinnitus loci were associated secondary to a role in causing hearing loss we compared the Manhattan Plots and preformed regional heritability analysis for the two traits (Figures 3 and 4). No shared significant loci were identified in the regional heritability analysis and other than at the *ZNF318* and *ARID5B* loci there is little overlap between the two Manhattan Plots. Furthermore, examination of the individual locus plot for rs4906228 upstream of RCOR1 in the hearing difficulty UKBB GWAS shows little evidence of an association with hearing loss (Figure 2) suggesting this association is not purely a secondary effect of an association with hearing loss. Locus plots for each of the 11 suggestive loci are displayed in Supplementary Figure 1.

**Figure 3.**
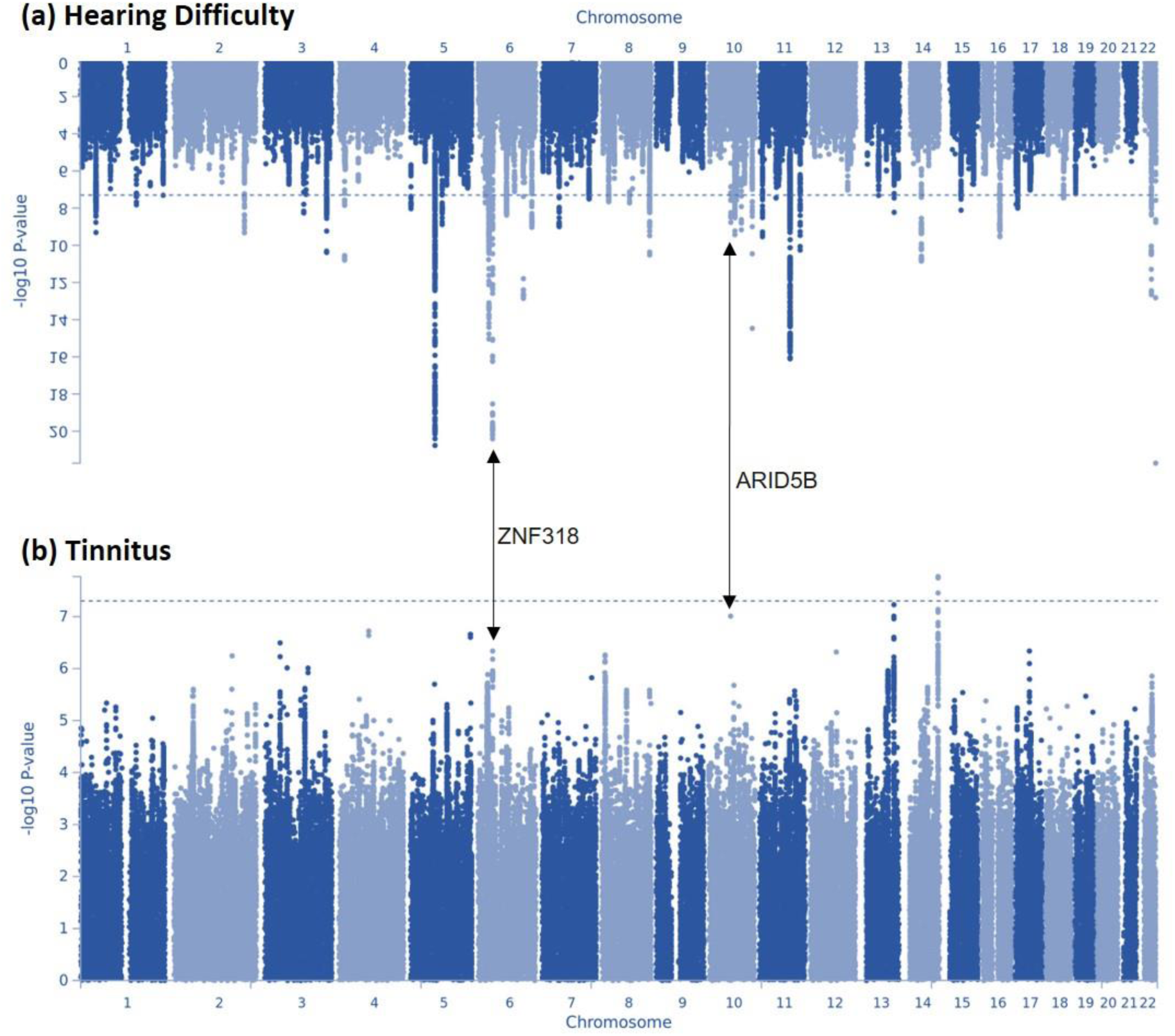
Manhattan plots displaying Hearing and Tinnitus GWAS results. (a) displays GWAS results for the hearing difficulty phenotype defined by Wells et al. 2019^22^ in the UK Biobank cohort, (b) displays GWAS results for the GWAS presented in this manuscript, with a tinnitus phenotype in the UK Biobank cohort. The Manhattan plots display the p values of all SNPs tested in discovery analyses. The threshold for genome wide significance (p < 5 × 10^-8^) is indicated by a dotted line. Loci common to both GWAS are annotated.

**Figure 4.**
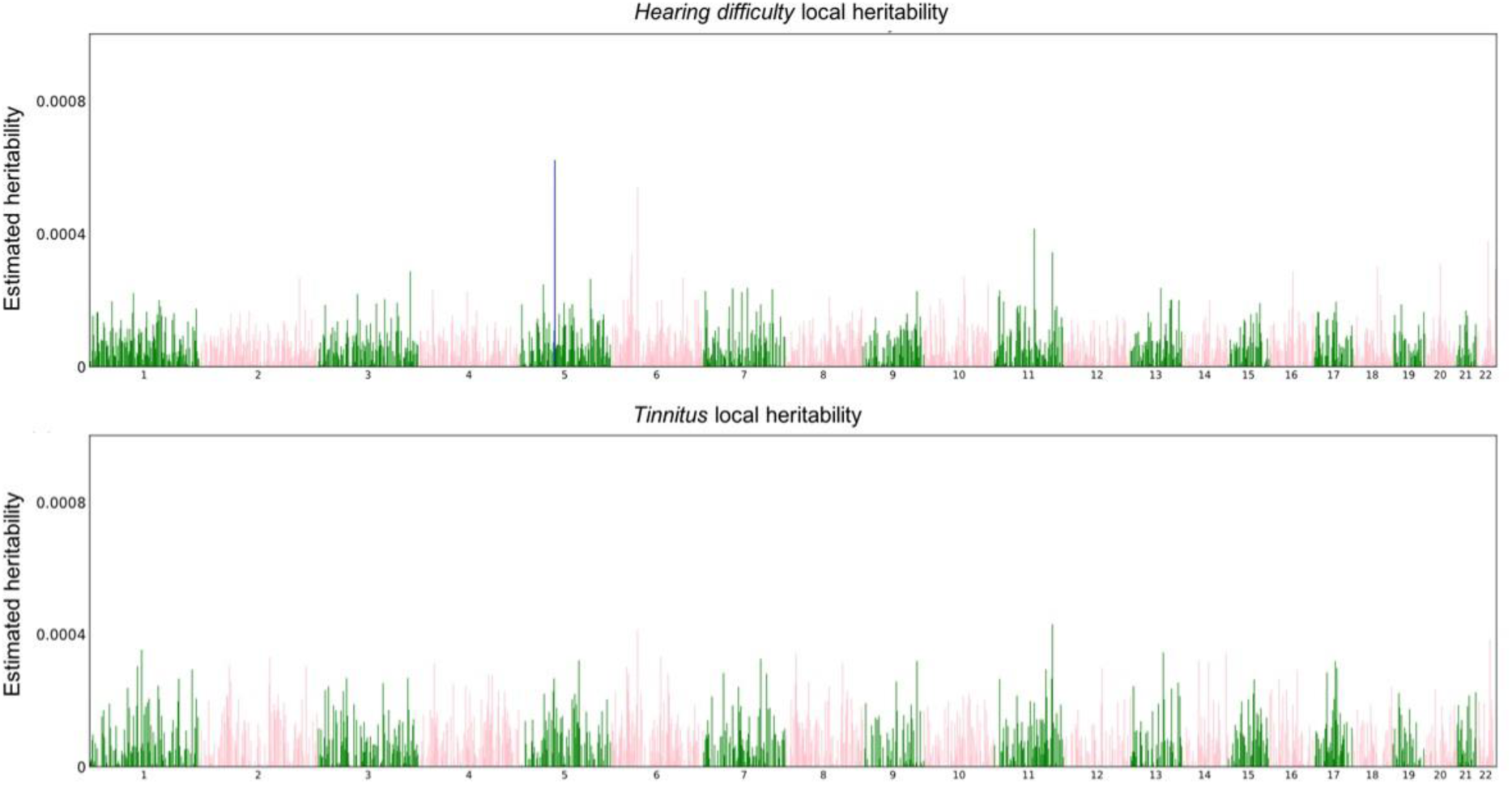
Manhattan-Style plots of regional heritability across the genome for hearing difficulty (top), and tinnitus (bottom). Estimated local SNP heritability for 1702 loci. Blue denotes an estimate of local_h2g = 0.0006, P<1.157E-05 on Chr 5 for hearing difficulty in the region between base positions 71240456-73759326. HESS total SNP heritability estimates were 0.1, SE = 0.005 for hearing difficulty and 0.0853, SE = 0.013 for tinnitus.

### Gene pathway and tissue enrichment analysis

In order to identify any properties or pathways common to the different tinnitus loci identified in the GWAS we undertook functional gene set enrichment analysis and annotation with genes mapped from SNPs associated at a suggestive level (P<1E-06) (Supplementary Table 2). Although a number of processes and pathways involved in auditory function were enriched at nominal significance, including cytoskeletal protein binding and regulation of anion transport, none of these are significant after Bonferroni correction for multiple testing. One gene enrichment finding which is significant after multiple testing correction is the presence of binding sites for the transcription factor hepatocyte nuclear factor 4 (HNF4) within six of the genes, *LMAN2, EXOSC7, ZDHHC3, SLC34A1, PDLIM7 and F12* (V$HNF4_DR1_Q3, Bonferroni corrected p value = 1.53E-03, Suppl Table 2).

### Gene Correlation Analysis

We used the linkage disequilibrium score (LDSC) to quantify any shared genetic variance between the genome-wide common genetic variants that influence tinnitus with other traits. LD Hub is a centralised database of summary-level GWAS results and interface for LDSC regression analysis including SNP heritability and genetic correlation^25^. We performed genetic correlation analysis between tinnitus and the 773 available traits on LD Hub. After removing the 2 traits that were used to create the tinnitus phenotype used in this study, 61 traits were significantly correlated with tinnitus (Supplementary Table 3). Of these, “Hearing Difficulty/Yes” and “Hearing Difficulty/ problems with background noise” were the most strongly correlated (rg=0.5173, p=5.36E-37 and rg=0.4285, p= 6.31E-26 respectively) confirming a strong genetic component shared between tinnitus and hearing loss. Other highly correlated traits (see Suppl. Table 3) included several involving recent experience of pain (rg=0.4039, p= 3.30E-18 for “Pain type(s) experienced in last month: Neck or shoulder pain”) and traits evidencing low mood or depression (rg=0.3196, p= 1.74E-16 for “Fed-up feelings”).

## Discussion

We have investigated genetic risk factors for a self-reported tinnitus by performing a GWAS in the UKBB with a sample size over 90,000 individuals, and identified 3 genome wide significant variant associations at the same locus. The lead SNP at this locus, rs4906228, is just over 8 kilobases upstream of the *RCOR1* gene in what Variant Effect Predictor^23^ describes as a regulatory region/promoter flanking region. Examination of the locus plot around rs4906228 (Figure 2) shows that the association with tinnitus, although strongest just upstream of the gene, spans the *RCOR1* coding region with 56 variants with p<E-06 over a 176 kilobase region along the length of the gene (Suppl. Table 1). RCOR1 potential role in tinnitus is interesting: while not previously linked to hearing or tinnitus it is a co-factor of RE1 Silencing Transcription Factor (REST) with which it forms a transcriptional repressor complex known to downregulate the expression of neuronal genes in non-neuronal cells through histone de-acetylases (HDACs)^24^. Dysregulation of the REST complex has been implicated in neurodegenerative disease including Alzheimer’s Disease^26^. A splicing mutation in *REST* has been reported as causing a progressive, non-syndromic, sensorineural hearing loss, DFNA27 in a North American family^27; 28^. Recently, Nakano et al. (2018)^27^ established the mechanism underlying deafness in this family. The sensory receptor cells in the inner ear *hair cells* inactivate REST using a cell specific splicing mechanism thereby allowing neuronal gene expression in hair cells. The DFNA27 mutation disrupts this alternative splicing mechanism resulting in repression of neuronal gene expression in hair cells, leading to hair cell death and deafness^27^. This suggests that there are distinct regulatory mechanisms and requirements for the RCOR1-REST repressor complex in both hair cells of the inner ear and neurons of the brain, both potential pathogenic sites for a tinnitus phenotype. We examined whether there was evidence that the association of *RCOR1* with tinnitus might be secondary to its effect on hearing by analysing any association with self-reported hearing difficulty in the UKBB cohort previous GWAS^22^. As Figure 2 shows there are no associations at P<5E-04 at this locus with hearing difficulty suggesting it is unlikely that the association with tinnitus is due to an increase in genetic risk of hearing loss.

Eleven other independent loci were associated with tinnitus at P<1E-6, providing a candidate gene list for further analysis. The association with *UBAC2* is just below the genome-wide significance threshold (p=5.90E-08) and lies in intron 2 of the gene. *UBAC2* encodes Ubiquitin-Associated Domain-Containing Protein 2 although there are a number of other genes that lie within this LD block including two G-protein coupled receptors *GPR18, GPR183*, and *FKSG9* which encodes Gasdermin-A, as well as 2 microRNAs (Suppl Figure 1). Three of these 11 suggestive associations are in or close to genes previously linked to hearing. The 2 base deletion 2:171146084_CTT_C is within an intron of *MYO3B*, encoding myosin IIIB, one of two myosin III isoforms that are responsible for the organisation and elongation of hair cell stereocilia that are critical for the detection of sound. Mutations in *MYO3A* underlie both dominant- and recessive forms of human hearing loss, DFNA73 and DFNB30 respectively^29; 30^. Furthermore, *Myo3a^-/-^Myo3b^-/-^* double knockout mice are profoundly deaf^31^. Two suggestive associations are downstream of genes that were previously identified as being associated with self-reported hearing difficulty in the UKBB cohort^22^, *ARID5B* and *ZNF318* although the lead SNPs associated with the two traits are different in both cases (see Table 3 and Figure 3). Both *ARID5B* and *ZNF318* encode transcription factors; AT-Rich Interaction Domain 5B and zinc finger protein 318. *ARID5B* has a role in regulating lipid metabolism, *ZNF318* is largely uncharacterized. It is therefore possible that the association of *MYO3B, ARID5B* and *ZNF318* with tinnitus is secondary to their role in hearing since tinnitus is usually manifested when there is a hearing loss present. However, it may be that the nature of the hearing loss caused by these genes variants creates a deficit which particularly potentiates the generation of tinnitus.

In order to investigate whether there are common functional properties or processes within genes associated with tinnitus that might pinpoint overlapping pathogenic mechanisms we performed gene set enrichment analysis. No enrichment of genes in specific molecular functions, biological processes, pathways or phenotypes was detected. However, HNF4 transcription factor binding sites were found to be significantly enriched within the gene set. HNF4 is a nuclear receptor and transcription factor, known to be critical in the regulation of hepatocyte development, nutrient transport and metabolism but it is also expressed outside the liver and thought to play a role in regulating gene expression relating to drug metabolism, lipid metabolism, cell proliferation, and inflammation^32^. Potential mechanisms connecting to tinnitus remain unknown but it is notable that Hnf4a^tm1b(EUCOMM)Hmgu^ knockout mice generated by the International Mouse Phenotyping Consortium are reported to have significantly reduced Acoustic Startle and Pre-pulse Inhibition response (PPI) (https://www.mousephenotype.org/data/genes/MGI:109128; accessed March 2020). Reduced PPI is often used as a surrogate indicator of tinnitus in animal behaviour experiments, although the validity of this has been questioned^33^. The lack of gene enrichment within the tinnitus GWAS gene set may well indicate the heterogeneity of tinnitus with multiple pathogenic mechanisms implicated.

Our study has identified a number of interesting candidate genes for further investigation, most notably *RCOR1*, but these require further replication and validation. The findings are limited by lack of replication group with comparable power in which to confirm these associations. The availability of such cohorts is an important barrier to further progress in tinnitus research. This is particularly pertinent to tinnitus because validation and characterisation in mutant mouse models is compromised by the lack of an established biomarker for the presence of tinnitus in animals. Tinnitus as defined in this study, that is *frequent* tinnitus (“*a lot, most or all of the time*”) has a heritability h^2^ of 0.30 which, although lower than some recent estimates in twin studies,^11; 12^ is still a sufficiently high genetic component to be tractable in future tinnitus GWAS if sufficiently large cohorts having the relevant phenotyping can be identified.

## Methods

### Participants and Phenotype Definition

The sample used for this study consisted of individuals who participated in the UKBB study. The UKBB is a national resource, initially set up to study lifestyle and genetic factors affecting ageing traits with the aim of understanding and improving healthy ageing at a population level. Over 500,000 volunteers attended 23 assessment centres across the UK between 2007-2013 where they donated samples for genotyping, completed lifestyle questionnaires and have standard measurements taken. The UKBB resource is described extensively elsewhere^19^.

Two questions regarding tinnitus were included in the UKBB ‘Health and medical history’ questionnaire that participants completed on touchscreen monitors while attending an assessment centre. The first question relates to symptoms of tinnitus and the second relates to the severity/nuisance of said symptoms. The first question is “Do you get or have you had noises (such as ringing or buzzing) in your head or in one or both ears that lasts for more than five minutes at a time?” to which participants could select one of the following responses: “Yes, now most or all of the time”, “Yes, now a lot of the time”, “Yes, now some of the time”, “Yes, but not now, but have in the past”, “No, never”, “Do not know”, “Prefer not to answer”. Participants responded to the second question; “How much do these noises worry, annoy or upset you when they are at their worst?” with either “Severely”, “Moderately”, “Slightly”, “Not at all”, “Do not know”, “Prefer not to answer”. The work in this study used data collected during the main recruitment phase only.

### Phenotype Definition

To derive a phenotype for association analysis, study participants were categorized using a case-control design based on responses to the question “Do you get or have you had noises (such as ringing or buzzing in your head or in one or both ears that lasts for more than five minutes at a time?” Participants that responded either “Yes, now most or all of the time” or “Yes, now a lot of the time” were assigned ‘cases’ and those that responded ‘No’ were assigned controls, described in Table 1. Participants that selected any of the additional responses were not included in the analysis. The cohort used for association analysis consisted of UKBB participants with ‘White British’ ancestry. The UKBB sample classification ‘White British’ is derived from both principal component (PC) analysis and self-declared ethnicity^34^. Samples with excess heterozygosity, excess relatedness and sex discrepancies were identified and removed prior to analysis, resulting in a sample size of n=91,424.

### Genotyping and Imputation

Two arrays were used to genotype the ~500,000 UK Biobank samples; 50,000 samples were genotyped on the Affymetrix UK BiLEVE Axiom array and ~450,000 samples were genotyped on the Affymetrix UK Biobank Axiom® array. The two arrays shared 95% of the >800,000 SNP genotype coverage. UKBB performed imputation centrally using the HRC reference panel and IMPUTE2^35^. Further SNPs that did not feature on this panel were imputed with the UK 10K and 1000G panel. Analysis presented here was conducted with version 3 of the UKBB imputed data, following QC performed centrally by UKBB, 487,409 samples were imputed and available for analysis.

### Association Analysis

Genetic association analysis was performed using BOLT-LMM v2.2 that uses a linear mixed-effects model approach to test between SNP dosages and the tinnitus phenotype. BOLT-LMM corrects for population stratification and within-sample relatedness. Additional adjustments were made for age, sex, UKBB genotyping platform and UKBB PCs1-10. Additional genotype QC included the implementation of a minor allele frequency threshold of 0.01 and INFO score >0.7 and removal of samples with a genotype call of <98%.

### Conditional and joint analysis

Conditional and joint SNP analysis was performed to identify independent signals within highly associated regions, using GCTA-COJO^21^. This analysis requires the linkage disequilibrium reference sample, which was obtained by random selection of 10,000 individuals from the UKBB cohort with White British ancestry. The reference sample size was selected to maximise power based on previous data simulations^21^. The distance assumed for complete linkage equilibrium was 10Mb and a cut off value of R2=0.9 was used to check for collinearity between the selected SNPs and those to be tested. Alleles with a frequency difference >0.2 between the reference sample and GWAS sample were excluded. Independent SNPs identified with GCTA-COJO were mapped to the nearest protein coding gene using variant effect predictor (VEP)^23^, genome build GRCh37. VEP was used to establish whether the SNP was in an exonic, intronic or intergenic region, and also the functional consequence of the variant at that position.

### Heritability Estimates

BOLT-LMM was used to calculate heritability (h2g) for the tinnitus phenotype as h2g = 0.105, SE=0.003. The estimate was then recalculated to the liability scale, using a case prevalence of 0.125. A region-based heritability estimation from summary statistics (HESS) was used on tinnitus and hearing difficulty trait from Wells et al. (2019)^22^ to partition genome-wide SNP heritability estimates into 1702 approximately independent loci. Partitioned heritability values for tinnitus, h2g = 0.105 and hearing difficulty, h2g = 0.35 were compared to investigate any loci sharing heritability percentage.

### Pathway analysis

Pathway-based analysis in GWAS is being widely used to discover multi-gene functional associations. Many gene set enrichment tools have been developed to test the enrichment of the associated genes in pathways, and ToppGene Suite^36^ is one of the sophisticated and easy to use tools suitable for this purpose. It comprises of many features such as pathway, Gene ontology, human and mouse phenotypes, diseases, drugs, etc to be included in enrichment analysis and it reported the enrichment p-values at both uncorrected and corrected levels. Enrichment of features at nominal level is useful for smaller number of genes input and for exploratory analysis. We prepared the gene list by mapping the lead SNPs from the Tinnitus GWAS to the nearest genes within 100kb of the gene transcription start/end site using Variant Effect Predictor as a positional mapping tool^23^. These were then entered into the web-interface ToppGene Suite tool as the gene list. The resulting enrichment features and values were analysed and evaluated for any interesting pathway involved with Tinnitus GWAS genes.

### LD Score Regression

The relationship between the test statistic and LD was studied (via univariate linkage disequilibrium score regression, LDSC), to calculate whether inflated test statistics are likely due to confounding bias, or the polygenic nature of the trait.

### Genetic Correlation Analysis

LD Hub is a centralised database of summary-level GWAS results and interface for LDSC regression analysis including SNP heritability and genetic correlation^25^. Here, we used LD Hub to analyse genetic correlations between tinnitus and all of the 771 other traits available from LD Hub. We set a multiple testing significance threshold of p<6.5E-5 (0.05/771)^25^.

## Data Availability

Summary Statistics will be made available on full publication on UK Biobank website and on request

http://biobank.ndph.ox.ac.uk/showcase/

## Acknowledgements

The research was carried out using the UK Biobank Resource under application number 11516. HRRW was funded by a PhD Studentship Grant, S44, from Action on Hearing Loss. The study was also supported by funding from NIHR UCLH BRC Deafness and Hearing Problems Theme. We would like to thank all the participants of UK Biobank.

